# A Deep Learning-Based Predictive Algorithm for Metabolic Syndrome Detection in the U.S. Population

**DOI:** 10.64898/2026.05.24.26354007

**Authors:** Cristina Correa, Ruben Solozabal, Ziad Akram Ali Hammouri, Fernando Gómez-Peralta, Hagai Rossman, Juan C. Vidal, David C. Klonoff, Eran Segal, Marcos Matabuena

## Abstract

**Objective:** To develop clinically operational, population-representative risk-score models for detecting metabolic syndrome (MetS) in U.S. adults by incorporating the NHANES survey design.

**Research Design and Methods:** We analyzed 36,812 U.S. adults from NHANES 1988– 2018. Seven models of increasing clinical complexity were trained and evaluated, ranging from basic demographics to full biochemical panels. We used a new deep-learning methodology for survey data with a predictive uncertainty quantification model.

**Results:** A model combining anthropometrics, vital signs and a basic lipid panel achieved an AUC of 0.923 at an estimated cost of 0.40€ per individual. Adding diabetes-specific biomarkers, including fasting plasma glucose (FPG) and glycated hemoglobin (HbA1c), yielded only marginal improvements.

**Conclusions:** This low-cost population-representative screening tool for MetS may help identify at-risk individuals and support data-driven public health interventions.

---

Metabolic syndrome (MetS) is a major public health concern worldwide [1]. Public health measures based on risk stratification tools are urgently needed, especially in primary care settings where complete diagnostic panels are rarely available at initial assessment. Here, we present a survey-weighted neural network approach with individual-level uncertainty quantification to predict MetS across models of varying clinical complexity. Whereas existing risk scores in this area have relied on logistic regression [2, 3], which assumes a linear relationship between independent variables and outcomes. Our method leverages the flexibility of neural networks to construct more sophisticated risk scores.

## RESEARCH DESIGN AND METHODS

We used the NHANES 1988-2018 dataset [4], that merges 614 individual files from NHANES III (1988–1994) and Continuous NHANES (1999–2018) into a unified analytical framework that covers 134,310 participants and 4,740 variables [5]. NHANES employs a stratified, multistage probability cluster sampling design to produce estimates representative of the non-institutionalized U.S. civilian population. Consequently, all analyses were performed using aggregated survey weights. Demographic data and clinical response data were merged using `participant IDs`. The analytical sample consisted of adults 18 years or older with complete data on all predictors and on the variables required to define MetS. The sample size was *N* = 36,812 participants.

### Metabolic syndrome definition

MetS was defined by consensus as the presence of three or more criteria of the definition of metabolic syndrome [6]: I. waist circumference >102 cm (men) and >88 cm (women); II. triglycerides ≥150 mg/dL; III. HDL cholesterol <40 mg/dL (men) and <50 mg/dL (women); IV. systolic blood pressure ≥130 mmHg or diastolic blood pressure ≥85 mmHg; V. fasting glucose ≥100 mg/dL.

### Neural network models

Following the survey-weighted neural network framework of [7], we trained seven multilayer perceptron (MLP) models with different combinations of clinical variables (Table 1). Comparing these models allowed us to evaluate trade-offs between clinical utility, measurement cost, model complexity, and predictive performance.

**Table 1:**
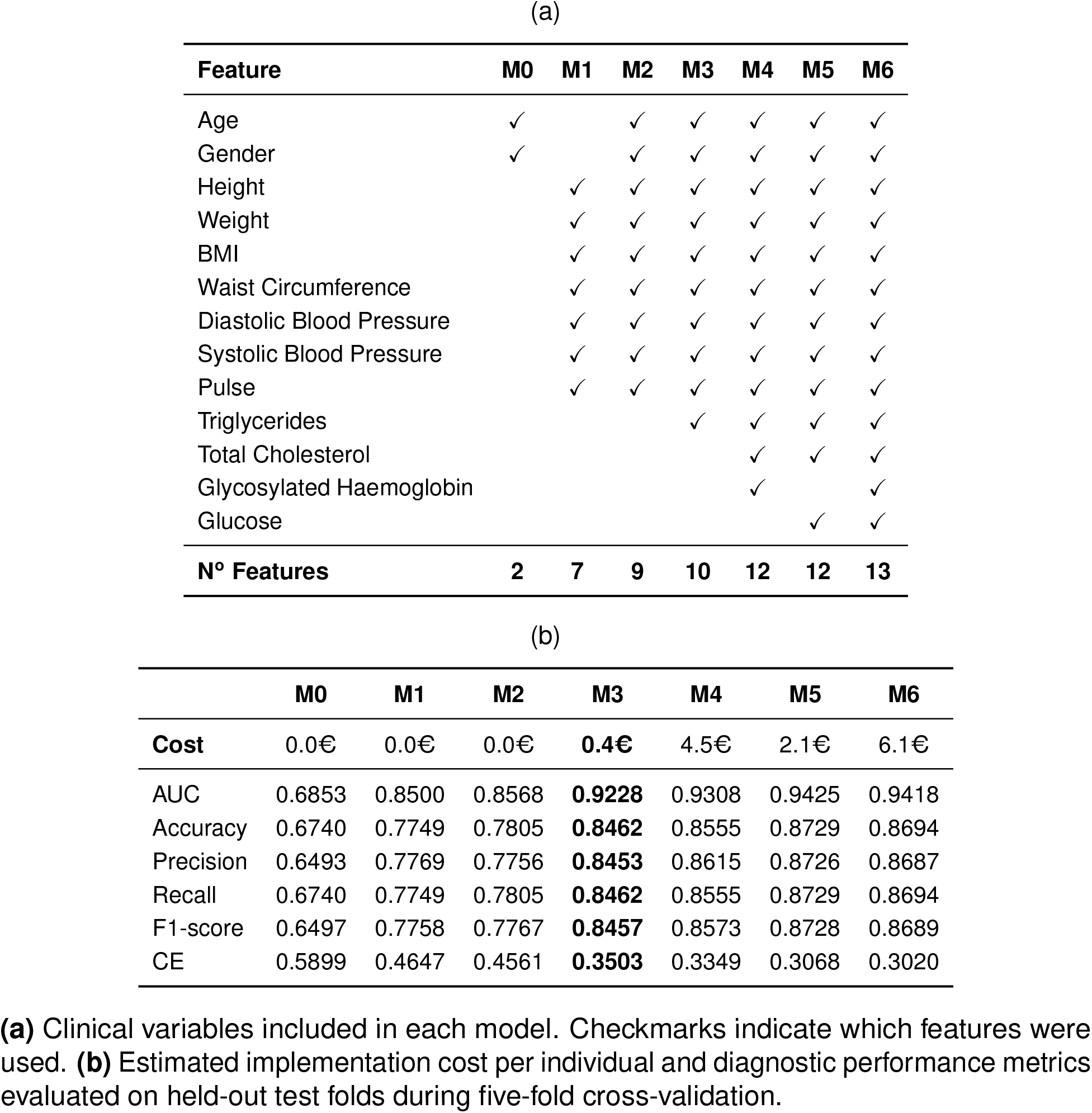
Performance comparison of model configurations on the test set.

Model 0 included only age and sex. Subsequent models gradually added anthropometric measures, blood pressure, and pulse (Model 1); demographic, anthropometric and vital signs variables (Model 2); triglycerides (Model 3); total cholesterol and glycated hemoglobin (HbA1c; Model 4); fasting plasma glucose (FPG; Model 5); and both HbA1c and FPG (Model 6). The models were optimized using the Adam optimizer and evaluated via five-fold cross-validation. All results were evaluated on held-out test folds during cross-validation.

The survey-weighted neural network method was implemented in Python. Implementation costs were expressed in cost units reflecting the estimated relative burden of obtaining the required clinical variables for each model, with specific cost estimates obtained from [7].

### Individual uncertainty quantification

Following the uncertainty quantification methodology described in [7], we calculated a conformal uncertainty score, *h*_*i*_, for each individual *i*. A larger *h*_*i*_ indicates greater model uncertainty for an individual, while a smaller value reflects a more deterministic prediction.

To understand which factors drive uncertainty in individual predictions, we fitted generalized additive models (GAMs) separately for each of the seven prediction models using the `mgcv` package in the statistical software R. GAMs are flexible regression models that can capture non-linear relationships. Here, continuous predictors were modeled as smooth terms *s*(*·*) via penalized regression splines, and sex was included as a linear term. All models were estimated using restricted maximum likelihood (REML). We focus the discussion on Model 3, because it represents the most clinically feasible option among the seven. This model captures most of the predictive signal using variables that are simple and inexpensive to obtain, while the incremental benefit of glycemic biomarkers is relatively modest compared to the additional implementation burden.

We fitted each GAM using both the original predictor scales and the standardized predictor scales. Models fitted on the original scale can directly assess statistical associations, whereas standardized models were used to compare the importance of each predictor.

### Model Performance metrics

The primary performance metric throughout this analysis is the AUC, which captures the model’s ability to discriminate between individuals with and without MetS. Complementary metrics are reported at the optimal threshold identified by the Youden index: accuracy (proportion of correctly classified individuals), precision (proportion of MetS predictions that are true positives), recall (proportion of actual MetS cases correctly identified) and F1-score (harmonic mean of precision and recall), all computed as macro-averaged estimates. Cross-entropy (CE) quantifies probabilistic calibration.

## RESULTS

### Predictive performance

Of the 36,812 adults in the analytical sample, 12,523 (34.0%) met criteria for MetS and 24,289 (66.0%) did not. Survey weights help mitigate class imbalance by upweighting or downweighting observations to better reflect population-level prevalence and reduce the distortion introduced into model training and evaluation.

As additional clinical variables were introduced, survey-weighted MLP performance increased monotonically (Table 1). Model 3 incorporated specific biochemical and anthropometric information into an 10-feature panel at an estimated cost of 0.40€, achieving an AUC 0.923, accuracy 84.6%, precision 84.5% and recall 84.6%. Applied to 1,000 adults with a 34.0% MetS prevalence, the M3 model would correctly classify approximately 846 individuals overall. If recall reflects MetS sensitivity, it would identify about 289 of the 340 true MetS cases and miss about 51.

The incremental gain from adding HbA1c (Model 4, AUC 0.931) or FPG (Model 5, AUC 0.943) was modest relative to the additional cost, 4.50€ and 2.10€ respectively; and the simultaneous inclusion of both biomarkers (Model 6, AUC 0.942) conferred no further improvement over Model 5 alone.

### Uncertainty quantification analysis

The goal of this analysis is to identify which clinical predictors are most strongly associated with greater individual-level model uncertainty, and to characterize the direction and linearity of those statistical associations. We focus on Model 3 because it offers a low-cost implementation without substantial loss in predictive capacity. Figure 1 displays the smooth GAM effects of each predictor on the conformal uncertainty index *h*_*i*_ for Model 3.

**Figure 1:**
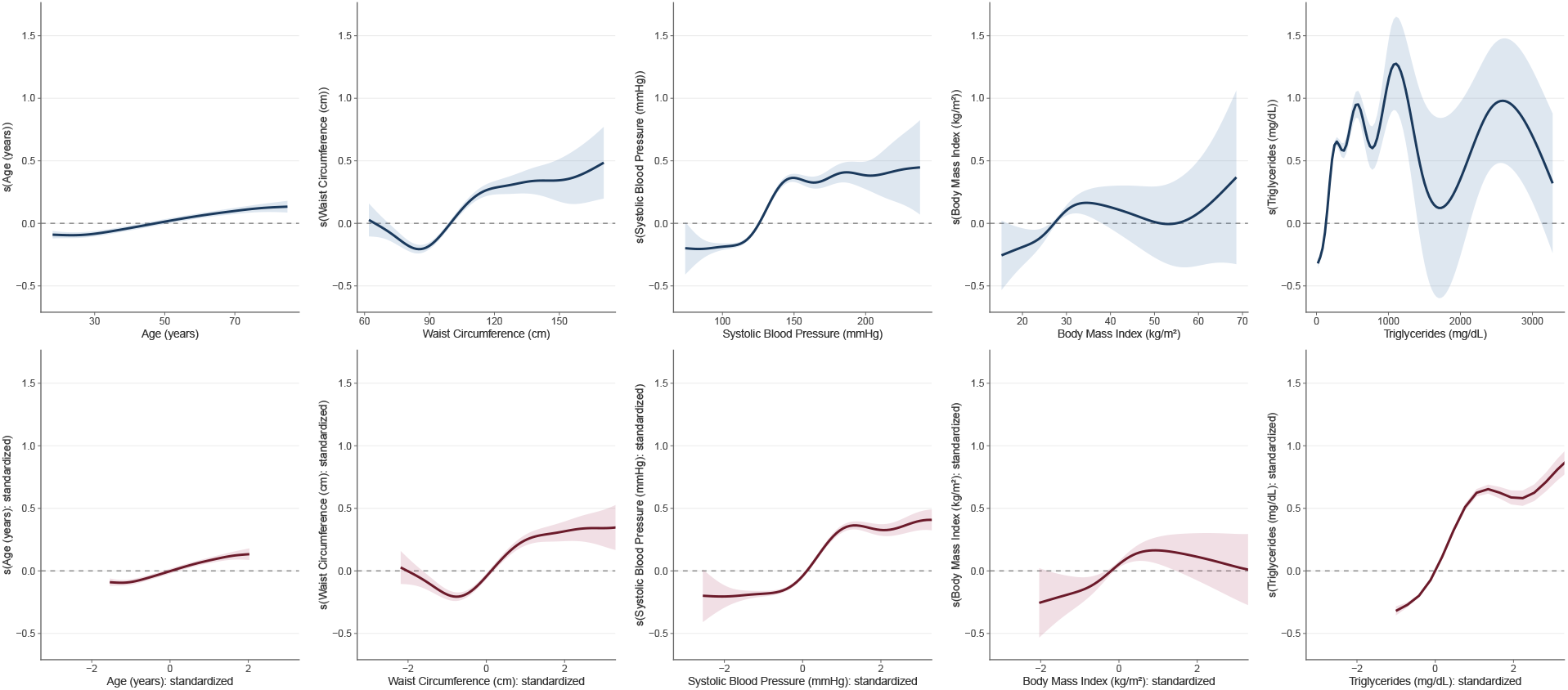
Association between clinical predictors and individual prediction uncertainty (*h*_*i*_) in Model 3. Results are shown on the original clinical scale (top row) and standardized scale (bottom row). Shaded areas represent 95% confidence intervals. Larger *h*_*i*_ values reflect greater model uncertainty for that individual.

Waist circumference, systolic blood pressure and triglycerides exhibited the greatest non-linear relationships with *h*_*i*_. The direction and shape of all associations were consistent across the standardized and original-scale representations, confirming their robustness to scale.

Taken together, these findings indicate that model uncertainty is greatest for metabolically borderline individuals. Individuals with obesity and a lipid profile that remains within or near the normal range represent the subgroup with the highest *h*_*i*_ as their anthropometric profile is suggestive of MetS, yet the absence of a clearly abnormal biochemical signal leaves the model in an indeterminate state. Conversely, individuals whose clinical variability falls into clearly pathological ranges, particularly those with high triglycerides or elevated systolic blood pressure, are classified with lower uncertainty, as their profile maps more deterministically onto the MetS phenotype.

## CONCLUSIONS

This study provides a set of survey-weighted neural network models for metabolic syndrome detection, designed to be representative of the U.S. population by accounting for the sampling design of NHANES. The framework further incorporates individual-level uncertainty quantification. It offers a clinically informative approach to population-level risk stratification surpassing conventional machine learning methods. In our discussion, we focus on Model 3 which represents the most operationally viable option for clinical implementation. It relies exclusively on demographics, anthropometrics, vital signs measures and triglycerides, but excludes HDL cholesterol, and diabetes-specific biomarkers, avoiding circular predictions. This feature set captures three of the five formal MetS diagnostic criteria that are easier to measure clinically. In the case of triglycerides, point-of-care measurements have shown high clinical agreement with laboratory measurements [8], making this feature set particularly well suited for deployment in low-resource settings and for low-cost screening.

The structure of prediction uncertainty is clinically coherent: the predictors most strongly modulating *h*_*i*_ are those entering the MetS diagnostic criteria. Their effects are largest in the borderline range of each threshold. This paradigm provides guidance on which factors drive model uncertainty, validating *h*_*i*_ as a triage signal capable of identifying metabolically ambiguous individuals for whom additional clinical evaluation would be most informative, and does not present risk assessment as a binary classification.

We acknowledge several limitations. First, the cross-sectional design precludes causal inference and longitudinal validation of the proposed risk stratification framework. Second, the AHA/NHLBI diagnostic thresholds adopted here are U.S.-specific and may not translate optimally across the full ethnic heterogeneity represented in NHANES [6], optimal cut-points for individual MetS components are known to vary with age, sex and ethnicity [9]. These personalizations might improve the phenotyping and final risk stratification of individuals. Nevertheless, this study presents important strengths: the use of NHANES survey weights ensures that findings are representative of the non-institutionalized U.S. civilian population, the large analytical sample (*N* = 36,812) provides robust findings, and the deep learning architecture captures complex non-linear relationships among predictors that conventional models may miss.

The models presented here can support decision-making by improving disease screening and healthcare operational capacity. Model 3, which relies on measurements routinely available in primary care settings, is especially suitable for deployment in low-resource environments, where scalable tools for metabolic syndrome screening are most needed.

## Data Availability

All data produced are available online at NHANES website

## Author Contributions

